# Implementation of flash glucose monitoring in four paediatric diabetes clinics – controlled before and after study to produce real world evidence of patient benefit. Statistical and health economic analysis plan

**DOI:** 10.1101/2022.11.22.22281186

**Authors:** R. Kandiyali, H. Taylor

## Abstract

This study seeks to answer two key questions:

- Does flash glucose monitoring improve outcomes in children with Type 1 diabetes compared to finger prick testing?
- What are the healthcare implications associated with flash monitoring when compared to finger prick testing in terms of staff time, experience and healthcare costs?

We report our combined Statistical and Health Economic Plan (SHEAP) for a controlled before and after study using routinely collected health record data on children who are eligible for flash glucose monitoring (Abbott FreeStyle Libre™) and a control population of children who continue to finger prick test in South West England. Our main analysis includes comparison of glucose control in terms of glycated haemoglobin (HbA1c) and UK health service costs relating to prescribing and loss of control (via record of diabetes related complications).

## Statistical and Health Economic Analysis Plan, SHEAP

## 1. Administrative Information

### FULL/LONG TITLE OF THE STUDY

Implementation of flash glucose monitoring in four paediatric diabetes clinics – controlled before and after study to produce real world evidence of patient benefit

### SHORT STUDY TITLE / ACRONYM

FLASH – Flash glucose monitoring in children with diabetes

### 1.2 Study registration number

REC: 21/WM/0195; IRAS project ID: 286988

### 1.3 SAP version 29^th^ July, 2022

### 1.4 Protocol version

**V**ersion 1.4 of the FLASH protocol

### 1.5 SAP revisions

*SHEAP revision history*

### 1.6 Roles and responsibility

Hazel Taylor, UHBW, Senior statistician

Rebecca Kandiyali, Warwick Medical School, Chief Investigator & Lead health-economist

## 2. Introduction

### 2.1 Background and Rationale for the Study

#### What is the problem being addressed?

Type I diabetes affects around 30,000 children and young people in England [1]. Management requires monitoring commonly by finger prick testing and injection of insulin multiple times a day. Automatic glucose monitoring devices greatly reduce the need for finger prick testing which is painful and has attendant issues in terms of anxiety, compliance and sleep disturbance.

Monitoring is essential for good glucose control. Poor control is associated with poorer outcomes including hypoglycaemia, diabetic ketoacidosis (DKA) and in the longer-term increased risk of micro (damaged small blood vessels) and macrovascular (damaged larger blood vessels) morbidity and mortality. There are different ways to monitor blood glucose; these include finger prick testing, flash monitoring and continuous glucose monitoring. Clinical teams encourage patients and families to make informed changes in their insulin dosing and dietary intake based on blood glucose levels, to improve overall diabetes control. For this study, we focus on whether flash monitoring offers improvements over and above finger prick testing, since continuous glucose monitors (which are more expensive) are not routinely available in the NHS for the majority of patients [2-4].

The only flash monitoring device available and authorised for use in the EU is FreeStyle Libre [5]. Libre measures glucose levels in the interstitial fluid via a sensor, worn on the upper arm continuously, containing a glucose sensing filament that sits under the skin. Each monitor lasts for two weeks. Readings are taken by scanning the sensor, via a Smart phone, or touchscreen reader device which can be read over the wearer’s clothing. As long as the sensor is ‘read’ within the last eight hours, additional data on glucose levels in the interval between readings is also downloadable. Additionally, parents and young people can download past readings (for instance all flash readings in the past month and the intervening estimates of glucose levels) from a cloud-based database for sharing with their clinical team. In this way, the history of flash readings can guide treatment decisions in clinic in addition to changes being made at home. Libre is generally welcomed by young people and parents as repeated daily finger prick testing is painful and disrupts daily activities and sleep. A small number of children may experience reactions (itching, skin redness) to the sensor and/or there may be other reasons why children and young people decline.

While Libre does not completely replace the need for all finger prick blood glucose testing, for instance where glucose levels are low or fluctuating rapidly, RCT evidence in adults suggests the need for finger prick testing may be reduced to a single strip every other day [6].

In November 2017, NICE recommended that Libre should be left to local CCGs to commission. This led to variable access to Libre for children, with some families choosing to self-fund and patient and stakeholder uproar. In November 2018 NHS England intervened to make Libre available from April 2019 on prescription to all eligible children and adults in order to remove the postcode lottery [7, 8]. Despite this move by NHS England, there is very limited real-world evidence that Libre improves glucose control and reduces diabetes related adverse events in children and young people. In addition, no study has specifically examined the short-term resource implications associated with the implementation of Libre. This is especially important, as NHS England states that the national funding arrangements for the Libre sensors are ‘time limited to include 2019/20 and 2020/21, which will allow time for CCGs and prescribers to implement NICE guidelines and recoup the financial benefits of (…) Libre’ [8]. The economic aspect of this study, the first of its kind, aims to provide NHS policy makers with information on whether recouping of costs is possible. This study will also provide evidence on the effectiveness, safety and perceived patient benefit of Libre.

#### Review of existing evidence – How does the existing literature support this proposal?

A scoping review of trial and observational evidence of Libre in children and evidence from regulatory and advisory bodies within the NHS, charities, and stakeholder groups has shown no randomised controlled trial in children and limited observational evidence.

#### Trial evidence

A single small RCT (n=60) in Israel is due to report in young people aged 12-17 this year [9].

In adults, RCT evidence (n=328) comparing Libre to finger prick testing demonstrated improvements in terms of: the primary outcome of reduction in time in hypoglycaemia (between group difference of 1.24 hours/day SE 0.239 p<0.0001 equating to a 38% reduction), reduced variability (glucose time in range) and a reduction in the number of blood glucose measurements in the short-medium term [10]. However, RCT evidence has not followed-up patients beyond 6 months [11].

#### Observational studies in children

In the UK, a single arm prospective study in 89 children found that Libre was 83.8% accurate when compared to blood glucose results, with a mean absolute relative difference of 13.9% between paired capillary and flash monitoring glucose levels[12].

A multi-centre European study [13] evaluated the use of Libre in 76 young people (baseline HbA1c 62.9 ±11.2; mean ± SD mmol/mol) aged 4-17 years for 8 weeks following a 2-week blinded baseline wear of the devices. Time in range (70-180 mg/dl) and HbA1c both significantly (p<0.01) improved compared to baseline [mean (±SD)] 0.9 (± 2.8) h/day increase in time in range and a -4.4 mmol/mol reduction in HbA1c to 58.5 mmol/mol. Scan frequencies were 12.9 / day and reduced the frequency of finger prick blood glucose measurements from 8.0 (baseline) to 1.0 daily. Satisfaction scores improved for parents and teenagers.

A multi-centre observational study in Israel, [14] found that self-funded regular (non-discontinuing n=59/71) users (children and young people aged 4-23 years) of Libre experienced significant reductions in HbA1c 8.86% (73mmol/mol) to 8.05% (64.5 mmol/mol)(p<0.01) at routine follow-up 6-12 months later. The study found no evidence of significant differences in time of glucose levels in range, reductions in hypoglycaemia or hyperglycaemia.

Preliminary evidence on parent and child outcomes is positive in terms of user acceptability, improved awareness, and better perceived self-management in terms of hypoglycaemia [12, 15, 16]

#### Economic evidence

Economic evidence in adults has been restricted to modelling and scenario analyses which demonstrate that Libre can be cost neutral or saving for people requiring frequent monitoring [6, 17]. Since most children and young people need to perform regular (8 times daily) tests, some of the cost of Libre sensors, which are replaced every 2 weeks, is offset by reductions in test strips needed for blood glucose measurements. Testing strips cost the NHS between £7 and £16 for a pack of 50 (11), which equates to a per patient spend of £8 to £18 per week, whereas the NHS tariff price of the Libre sensor is £35 every two weeks [18]. Economic evidence to date has ignored both the staffing resource required to review glucose measurements and the potential costs and benefits of long-term glucose control [6, 17] so studies assessing both the real world costs of implementation as well as the cost implications over time are needed [19].

##### RATIONALE

#### Why is this research important in terms of improving the health and/or wellbeing of the public and/or to patients and health care services?

Type 1 diabetes is a demanding lifelong condition. Children, young people, and their caregivers need to have the knowledge, skills, and confidence to achieve good glucose control.

Children and young people with poor glucose control have higher incidence of developing long-term complications later in life such as kidney failure requiring dialysis or transplant, limb amputations, blindness and cardiac problems leading to early death [20]. Additionally, in the short-term there are the risks of emergency admissions associated with hypoglycaemia and DKA. The NHS spends £1 billion annually on type 1 (T1) diabetes of which around 80% goes on treating complications [21].

Libre offers an alternative and potentially inexpensive method of glucose monitoring that avoids painful finger prick testing. While Libre has been welcomed by clinicians and patients, the impact of implementation on health care provision has not been studied. From talking to both the South West Children and Young People (CYP) Diabetes Research Network and young people and their parents, one issue with the national roll-out of wearable technologies is of information overload.

The convenience of Libre compared to finger prick testing has been shown to increase the frequency of readings, providing families and the clinical team with much more information. This means that families may become aware of fluctuations which were not picked up by less regular finger prick testing. The reality is that blood glucose levels are highly individual and may fluctuate widely depending on diet, activity, illness, and basal (background) insulin so that keeping ‘in range’ (4-10 mmol/l for short-term blood glucose control) is challenging [2].

This variation may cause concern in parents which needs to be balanced against the comfort, convenience and reassurance that Libre may provide. However, the initial parent and young people involvement work we have done suggests that the reduction in disruption to activities and sleep compared to finger prick testing is highly valued. One parent told us that it can be difficult to maintain regular blood glucose measurements when her nine-year-old is playing with her friends with the daughter repeating “Why me?” which is distressing for both mother and daughter. Meanwhile Diabetes nurses and diabetologists have told us that they are not “analysts” and that they worry that interpretation of data is taking them away from their caregiving roles. The young person we spoke to using Libre (adolescent male, parent self-funding Libre prior to April ‘19) said he felt confident about using the device but found the way the download visualises recent readings “a bit mathsy” and something “I don’t really look at…the graphs are for my doctor”.

Guidance from NICE acknowledges that the resource implications of Libre are “uncertain” [11]. If adopted, it is likely that the additional costs of the sensors and readers may be partially or even wholly offset by savings in the number of blood glucose testing strips but there are other implications such as clinical time spent interpreting glucose profiles. In the longer-term, costs may be saved if Libre leads to better monitoring and control with fewer admissions for diabetes related complications.

The idea for this study came from the PI’s work within the local CCG, with the study designed as a response to provide a solution to the evidence gap surrounding flash monitoring. Our study fits with the vision of the Royal College of Paediatrics and Child Health (RCPCH) response to the NHS Long Term Plan in terms of placing children at its heart and ensuring that children with long-term health problems have access to personal and tailored care across their life-course and through transition to adult services [20]. In addition, the findings will be relevant to NHS England and clinicians and others involved in the provision and commissioning of Diabetes care. Using real world routinely collected data is also efficient and allows us to establish patient benefits and costs in a relatively short time period. We have included co-applicants and collaborators linked to the policy, commissioning and delivery of diabetes services to expedite pathways to impact.

### 2.2 Aims and Objectives

The overarching aim will be to establish guidance which will improve the provision of care for young people with diabetes and their families. The purpose of the guidance will be to provide an assessment of the value of Libre in this patient group along with practical information to patients and healthcare staff in relation to use of Libre.

#### Objectives

i. To compare long-term blood glucose control (HbA1c) and acute diabetes complications in paediatric patients before and after they start using Libre, and with a contemporaneous comparator group who continue to finger prick test.
ii. To compare the NHS secondary healthcare resource and costs in paediatric patients before and after they start using Libre.
iii. To identify the benefits, challenges and burden to NHS staff of using Libre in their care provision.
iv. To ask young people and parents about their experiences of using Libre.
v. To produce guidance on the use of Libre which we can disseminate regionally and nationally.

## 3. Study Design

### 3.1 Study Design

This study will employ a multi-method approach to draw together clinical, economic and qualitative evidence. The study plans to use observational cohort data from implementation in practice using observational data from up to 4 study sites (Bristol, Exeter, Swindon and Plymouth).

Data will be collected from a cohort of patients who have Libre initiated at any time between April 2019 and June2020. Data will also be collected on patients at the 4 study sites who continued to finger prick test despite being eligible for Libre. These patients will be a control group.

Routinely collected data which are held in local hospital databases will be used and will include data relating (up to) 12 months prior to Libre being initiated and 12 months after. These data are collected for the purposes of national audit and are held in centre databases. While the majority of the data is held on a local centre database; data on Libre is held on the Libreview platform and accessed via a trust login. Data will be downloaded and shared in a secure and anonymised format by the senior diabetes research nurse from the paediatric diabetes clinic databases to the study statistician/health economist. No identifiable data will be held by the research team.

### 3.2 Sample size

For adults and children, CCGs anticipate that 20% of T1 diabetes patients will be eligible for Libre, but this is contentious, with arguments mounting that criteria are restricting access in those with need [22]. Our network diabetologists anticipate that 30-40% of the combined population will be eligible for Libre as nearly all primary school aged children (about 27% of the T1 diabetes paediatric caseload) will need help monitoring. Given the combined population of young people with T1 diabetes (aged 0-19) across our 4 centres who manage their diabetes by multiple daily injections in 2017-18 and so would be eligible for Libre was 778, a sample size of 200 would need approximately 25% of those eligible to take up the libre option within the study period.

The primary analysis of the primary outcome will be a paired t-test to compare the mean difference in mean HbA1c in the before and after periods. A sample size of 200 will have 90% power to detect an effect size of 0.230 (equivalent to being able to detect a mean of paired HbA1c differences of 1.4 mmol/mol, assuming a standard deviation of the differences of 5.9 mmol/mol [13]) with a significance level of 0.05 using a two-sided paired t-test. Clinical significance will be based on a minimally clinical important difference of 5 mmol/mol.

Note that all eligible patients, using Libre or fingerprick testing will be included in the sample and the analysis, even if this exceeds 200, subject to cost and time constraints.

### 3.3 Framework

All hypothesis testing will test for superiority.

### 3.4 Statistical interim analyses and stopping guidance

No formal interim analysis is planned.

As this study uses routinely available observational retrospective data, the study won’t be stopped.

### 3.5 Timing of final analysis

The final statistical analysis will take place only once all data has been collected and entered onto the database.

### 3.6 Timing of outcome assessments

All outcome data will be collected for patients who started Libre between April 2019 and June 2020. Data will be collected for each 3 month period during the year prior to starting Libre, and for the year after starting Libre. Although it is hoped that it will be possible to collect data for the year prior to starting Libre for all Libre patients, it is recognised that some patients now start Libre quickly after being diagnosed with Type 1 diabetes, and so for these patients there will not be a year’s worth of pre-Libre data available. However patients will only be included if is it possible to collect a minimum of 3 months worth of data prior to starting Libre.

Outcome data collected will include:

- Glycated haemoglobin (HbA1c)
- Serious hypoglycaemic events requiring attendance at hospital
- Hospital admissions for Diabetic Ketoacidosis
- Other diabetes related hospital admissions (e.g. for stabilisation)
- Diabetes related outpatient consultations
- Mean frequency of blood glucose finger prick tests for each 3-month period
- Average blood glucose over 2 weeks in mmol/l (where the patient has achieved a mean daily fingerprick frequency of 4 or more)

For patients on Libre, the % of time glucose in 4-7 mmol/l range and the mean frequency of scans for each 3-month period will also be collected.

For the control group of patients, who are continuing to finger-prick test, as there is no implementation date to determine the period of data collection, a sham implementation date will be created, and data will be collected for up to a year (minimum 3 months) prior to this sham implementation date, and for a year after.

The allocation of the sham implementation date will be carried out by the study lead who is blinded to personal identifiable information. The distribution of Libre dates will be used to form a distribution of sham dates for each centre which will then be randomly assigned to control participants (i.e. stratified random selection).

## 4. Statistical Principles

### 4.1 Confidence intervals and P values

All p values will be 2 sided, and the significance set at 5% level.

All confidence intervals will be 95% confidence intervals.

### 4.2 Adherence to Libre

The number of patients who withdrew from Libre and went back to finger-prick testing within the 12 month follow-up period will be reported.

### 4.3 Analysis Populations

The primary analysis will include all patients who started on Libre, even if they stopped using Libre in the 12 month follow-up period. A secondary analysis will only include patients who continued on Libre for the duration of the 12 month follow-up.

## 5. Study Population

### 5.1 Screening Data

Individual sites will run searches to identify patients who were prescribed Libre between April 19 and June 20 and those who are eligible controls (primarily those on multiple daily injections and a small number of patients on pump therapy without CGM).When undertaking screening of potentially eligible patients from these lists, the direct care staff doing the extraction will keep a record of any patients that do not fit the eligibility criteria.

### 5.2 Eligibility

- Eligible for Freestyle Libre (18 years of age or younger as of 1st January 2021 and requiring monitoring >8 times a day as indicated by clinical review) [8]
- Diagnosed with T1 diabetes^1^
- Has data on finger prick blood testing (controls) or Libre for at least 12 months*
- Not on continuous glucose monitoring with integrated insulin pump during the applicable study period. (Note the study period varies depending on the date of initiation of FreeStyle Libre. Data collection will be retrospective and limited to April 18 – July 2021^2^).

To be eligible for the Libre group, Libre needs to have been initiated at any time between April 2019 and June 2020.

Given that this study was designed to look at the implementation of the first generation Libre (not Libre 2) we also made an assumption that **all (control or fingerprick) patients would be at least 3 months from diagnosis** (and therefore able to provide at least 3 months of pre-Libre fingerprick data.

### 5.3 Recruitment and Consent

As this study uses anonymised retrospective data collected from routine clinic databases, we have permission from HRA to run a study without consent. We will follow STROBE guidelines and report a breakdown of the number of patients eligible and the number of patients entered.

### 5.4 Withdrawal/follow-up

Withdrawals are not possible as the anonymised routine clinic data collected does not require participant consent. There may be a small number of patients for which data is lost, as they move house during study period, however patients need to have at least 12 months of data to be included.

It is possible there may be some missing data at the time of the first lockdown, due to the lack of clinics and hence data collection (see 6.3).

### 5.5 Baseline characteristics

Patient baseline data will be collected from routine database records and will be tabulated for the Libre group and the control group to give an overview of the two groups. Group summaries will use percentages for binary variables and either means/SDs or median/IQR for continuous variables according to their distributional properties. As this is an observational study, statistical testing, will be used to identify baseline differences between the groups.

Baseline data will include: sex, height, weight, partial date of birth (year and month) to calculate age, duration of diabetes, date commenced/stopped Libre to calculate length of time on Libre, whether Libre 2 was initiated, type of insulin therapy (e.g. basal/bolus regimes), concomitant pump therapy, comorbidities (coeliac disease and hypothyroidism in particular) and ethnicity. Area-level deprivation will be calculated (from full postcode although the latter will not be stored).

Data on service characteristics will be collected and reported for each centre, and will include: size of the centre as determined by number of young people with T1 diabetes on list; consultant and nurse to patient ratios, and type of training offered to new users of Libre at each centre.

### 5.6 Potential Confounding Covariates

It is recognised that some patient characteristics could affect whether or not a patient decides to use Libre. As part of the statistical analysis, there is a plan to compare HbA1c in Libre and non-Libre users, and these patient characteristics could be potential confounding factors in this analysis.

The potential confounding variables include those listed in the previous section.Time varying factors that will also be considered are change to Libre 2 and the impact of covid 19 (which for some quarters meant fewer clinic appointments, more missing data, and different HbA1c measurement methods).

It will be possible to take account of these factors in the statistical analysis, by adjusting for them in a mixed effects model, comparing the HbA1c in Libre users and non-users.

## 6 Analysis

### 6.1 Outcome definitions

The primary outcome is overall glycaemic control to be measured in terms of glycated haemoglobin^3^ (HbA1c) measured in mmol/l and will be captured for up to 12 months before and 12 months after the child starts using Libre. More recently, some children have moved from finger prick testing to Libre quite quickly, so these children, may only have one quarters worth of data prior to starting Libre (See Eligibility criteria footnote 1). Children and young people typically have follow-up clinics every 3 months, and so the aim is to have between 1 and 4 pre and 4 follow-up time points with HbA1c readings for each patient. The mean HbA1c before and after Libre is started will be calculated from these readings and used as the primary outcome, even if there is only one reading available.

For the control group, not using Libre, a sham implementation date will be generated, and data will be collected using 4 pre and 4 follow-up time points from this sham date in a similar way.

Secondary outcomes will also be captured for up to 12 months before and 12 months after the child starts using Libre or from the sham date for those in the control group. Secondary outcomes include:

- Serious hypoglycaemic events requiring attendance at hospital.
- Diabetic Ketoacidosis hospital admissions.
- Other diabetes related hospital admissions (e.g. for stabilisation).
- Diabetes related outpatient consultations.
- Percentage time glucose in 4-10 mmol/l range (only available for users of Libre). (This is automatically calculated within the Libreview platform and available for download in clinics).

Process outcomes

- Mean frequency of scans for each 3-month period (Libre users only)
- Mean frequency of blood glucose finger prick tests for each 3-month period.

### 6.2 Analysis methods

#### Primary Outcome (HbA1c)

The primary analysis will include all patients who were initiated on Libre in 2019/20, regardless of whether or not they continued on it. A secondary analysis will only include patients who continued on Libre for the duration. A paired t-test will be used to compare the mean HbA1c prior to LIbre with the mean HbA1c post Libre.

Subgroup analyses by age and sexfor the outcome HbA1c may be undertaken. The age categories for the subgroup analysis, will depend upon the numbers, but it is anticipated that the following categories will be used; 8-11, 12-15, 16-18.

The analysis of the primary outcome assumes that the appropriate distributional assumptions are met. It is likely that the HbA1c will need to be log transformed for the statistical analysis as HbA1c has been shown to have a log normal distribution.

#### More detailed Analysis of HbA1c

A further more detailed analysis making full use of the HbA1c data will follow, which will include comparison with the control group, and adjustment for confounding variables. As this is an observational study, the analysis is likely to evolve and be data led.

Time of the HbA1c reading (in days) will be calculated, by calculating the days between the date of the HbA1c reading and the date of libre implementation for the libre users, and between the date of the HbA1c reading and the sham date for the control group. To enable box plots and summary statistics to be produced, this time between the reading and libre implementation/sham date, will also be categorised into 4 quarters pre and post libre implementation/sham date.

Box plots showing the median readings in each of the 4 quarters pre and post Libre, for the Libre and control group will be produced, to illustrate any trends over time.

In order to further understand the relationship between the potential confounding variables (such as age, sex, ethnicity, time since diagnosis) and HbA1c over time, box plots will be produced by these groups.A further plot will separate out the libre 2 users. In order to see how the covid 19 pandemic impacted HbA1c, a box plot will be produced with date of quarterly reading (i.e April – June 20, July – September 20) on the x axis, rather than quarter number pre or post libre implementation/sham date.

Initial modelling will be carried out to explore the longitudinal nature of these relationships with HbA1c. This analysis may inform, how to best use the potential confounding variables in the multilevel analysis.

A mixed model will be used to compare HbA1c in those using Libre and those eligible but not using Libre, using data from as many time points as available. The log of HbA1c is likely to be used as the outcome, as it is standard to log transform HbA1c in statistical analysis. Data checks will made to check that using the log transform is appropriate in this instance

The model will include a dummy variable to flag whether the HbA1c values are pre or post Libre values, in addition to a variable to flag whether or not the patient is in the Libre group or control group. (Note for the control group, the term post Libre, refers to values from after the sham implementation date). Patient characteristics which are likely to be acting as possible confounders will be considered for the model as fixed effects; namely, deprivation, ethnicity, pump therapy, sex, duration of diabetes, age, height and weight at implementation/sham date.Centre will be included as a random effect. Measurement method for HbA1c (as the HbA1c tended to be measured in the laboratory during some of the covid period which gave a higher reading) and change to Libre 2 will be considered as time varying confounders. Time (pre post libre implementation/sham date) will be considered inthe model as a continuous variable. All variables listed will be considered for the mixed model and included if they seem important.,

The modelling approach will be data led and informed by the box plots.but is likely that a final model will be informed by a series of simple models. These simple models are likely to only consider sections of the dataset. From the final model, the mean difference with 95% confidence interval in HbA1c pre and post Libre will be presented. Simple models which may considered are:

A mixed model for just the Libre group, which may include fitting an interaction between time and pre/post implementation date.. (This could also be repeated for the control group).

A mixed model comparing the Libre and control group prior to Libre implementation. An interaction could be fitted to inform whether the relationship between HbA1c and time, differs between the libre and control groups pre implementation

It is expected that it will take at least 3 months for HbA1c readings to settle following the introduction of Libre. As a result of this, modelling for the Libre group, may exclude data from the first 3 months post implementation.

A mixed model including Libre users and controls, using the mean log HbA1c reading post implementation/sham date for the outcome (as with the paired t-test), adjusting for the mean log HbA1c reading pre-implmentation/sham date.

A mixed model using all individual HbA1c readings, for Libre users and controls, pre and post implementation / sham date.

Two sets of models will be produced. The main analysis will include all patients who were initiated on Libre in 2019/20, regardless of whether or not they continued on it. A secondary analysis will only include patients who continued on Libre for the duration in the Libre group.

#### Analysis of Secondary Outcomes

The percentage time blood glucose is in 4-10 mmol/l range for each 3 month period is only available for patients who have Libre. Box Plots showing the percentage time in range for each quarter will be produced. The percentage of patients meeting the target figure of 70% of time in range, will be reported by quarter with 95% confidence interval.. The percentage time blood glucose is in range will also be plotted against HbA1c.

The number of patients experiencing the secondary outcomes (serious hypoglycaemic events requiring hospital attendance, diabetes ketoacidosis hospital admissions, other diabetes related hospital admissions or extra diabetes outpatient consultations) in the 12 months before and after Libre will be reported. As it is likely that a small number of patients may experience the outcome more than once, and for some patients there will be before data for less than 12 months, this data will be reported as incident rates, for pre and post, and for the Libre and control group (See Draft Table). Incident rates will be presented as number of events observed over the total number of person-years of observation. Differences between the rates will be reported by calculating differences and 95% confidence intervals

#### Analysis of Process Outcomes

Summary statistics will be reported for the process outcomes, the mean frequency of scans for each 3 month period ((Libre users only), and for the mean frequency of blood finger prick tests for both groups. Box plots will also be produced.

### 6.3 Missing Data

Due to lockdown during the COVID 19 pandemic there is likely to be some missing data. There will also be some missing data, due to some patients in the Libre group having less than 12 months worth of data available prior to starting Libre.

The nature of the missing data will be explored, and appropriate methods to deal with the missing data will depend upon the extent and the nature of the problem.

The mixed effects multilevel models to be fitted, can handle missing outcome data, and will still be able to include patients, who do not have outcome data for all timepoints available.

### 6.4 Additional Analyses

No additional analyses are anticipated however as this is an observational study, the final statistical analysis will be dependent upon the data observed, and so will evolve.

### 6.5 Statistical software

All data will be analysed in Stata (version to be confirmed).

## 7 Health Economic Analysis

### Overview

We will value resource data associated with loss of glucose control (i.e. hypoglycaemic, DKA and other diabetes complications as listed in 6.1). We will also collect information on the provision of ‘structured training’(4) associated with Libre in terms of staff and patient time and any non-staff resources, with further resource implications of training (e.g. staff capacity to provide training) probed as part of the focus groups/staff interviews described in the study protocol. We propose to examine resource and costs from an NHS secondary care perspective for the before and after periods for both groups of patients i.e. those who do, and do not take up Libre between April 2019 and June 2020).

### Clinic data

Secondary care prescribing of blood glucose monitors, testing strips, Libre sensors and readers may not be available from the centre database; resource use may be estimated from data obtained via application to a single clinical commissioning group and valued using NHS tariffs [18]. The cost of treating hypoglycaemic, DKA and other diabetes complications including ED attendances will be calculated by assigning the appropriate healthcare resource group (HRG) and associated reference cost for the episode (Table 1). The choice of framework for the economic analysis will be guided by input from CCG and other decision-makers (NHS England, NIHR, hospital providers, CYP Diabetes Research networks). For the report and main study manuscript, the economic analysis will be restricted to a cost comparison (Table 2). A sensitivity analysis will adjust for the effects of possible confounders (adjustment will be guided by inspection of clinical results as described in the SAP); elsewhere methods of good practice for economic evaluation of observational data will be followed [23-25].

### Dissemination to commissioners

Preliminary discussions have suggested that the metric for cost-effectiveness may include net cost to the CCG, cost/event avoided and budget impact modelling. Follow-on funding may be sought by separate application for economic modelling to capture the long-term costs and consequences of Libre. The analysis of both glucose control (Methods 1) and health service resource and costs (Methods 2) will provide commissioners and NHS England with information about the budget impact and potential health benefits of Libre for the T1 diabetes paediatric population.

### Non-participant observations of clinic time

We will use structured non-participant observations guided by time and motion study methodology (up to n=20) of consultant-led diabetes clinic appointments to identify the healthcare staff resource (time/activity) used to manage patients associated with specific tasks using Libre in clinics (up to n=10) as compared to managing patients who continue with finger prick testing (up to n=10).

As this element is exploratory (in that it is intended to provide complementary evidence to the qualitative staff interviews surrounding the issue of data overload associated with the implementation of FreeStyle Libre), data will be presented in terms of descriptive statistics (frequency of activity and mean times) supplemented by commentary based on the observer’s experience.

## Data Availability

Not applicable for this article

## Notes on eligibility

1 For newly diagnosed patients, we require 15 months of data

2 An extra month is added here, to include patients who might have had a clinic slightly beyond 1 year following June 2020.

* this implies a total data collection period of up to 24 months

## References

1. National Paediatric Diabetes Audit 2017/18, Available at: http://npda-results.rcpch.ac.uk/annual-reports.aspx. Accessed 14/05/2019. 2019.

2. NICE, NICE guideline [NG18]. Diabetes (type 1 and type 2) in children and young people: diagnosis and management. 2016.

3. NICE, Diabetes in children and young people. Quality statement 4: Continuous glucose monitoring in type 1 diabetes. Available at: https://www.nice.org.uk/guidance/qs125/chapter/Quality-statement-4-Continuous-glucose-monitoring-in-type-1-diabetes. Date accessed 04/06/2019. 2016.

4. Diabetes UK, Diabetes UK Consensus Guideline for Flash Glucose Monitoring. Available from: https://www.diabetes.org.uk/resources-s3/2017-09/1190_Flash%20glucose%20monitoring%20guideline_SB_V9[4].pdf?_ga=2.137083376.1339632840.1505301182-2056973880.1505301182. Accessed 04/06/2019. August 2017.

5. Abbot laboratories. https://www.freestylelibre.co.uk/libre/. Accessed 04/06/2019. 2019.

6. Healthcare Improvement Scotland. Evidence Note, Number 81. What is the clinical and cost effectiveness of Freestyle Libre flash glucose monitoring for patients with diabetes mellitus treated with intensive insulin therapy? Available at: http://www.healthcareimprovementscotland.org/our_work/technologies_and_medicines/topics_assessed/shtg_009-18.aspx. Accessed 19/07/2019. July 2018.

7. NHS England, NHS to provide life changing glucose monitors for Type 1 diabetes patients. Webpage Available at: https://www.england.nhs.uk/2018/11/nhs-to-provide-life-changing-glucose-monitors-for-type-1-diabetes-patients/. Accessed 14/05/19. 2018.

8. NHS England, Flash Glucose Monitoring: National arrangements for funding of relevant diabetes patients. Available at: https://www.england.nhs.uk/publication/flash-glucose-monitoring-national-arrangements-for-funding-of-relevant-diabetes-patients/. Accessed on 04/06/2019.

9. ClinicalTrials.gov, ClinicialTrials.gov identifier: NCT02776007. Patients Perceptions of Using the “Libre” System Compared With Conventional SMBG in Adolescents With Type 1 Diabetes The Libre Sat Trial. Details at: https://clinicaltrials.gov/ct2/show/NCT02776007. Date of access: 25/06/2019.

10. Bolinder, J., et al., Novel glucose-sensing technology and hypoglycaemia in type 1 diabetes: a multicentre, non-masked, randomised controlled trial. Lancet, 2016. 388(10057): p. 2254–2263.

11. NICE, Freestyle Libre for glucose monitoring. Medtech innovation briefing. Available at: https://www.nice.org.uk/advice/mib110. Accessed: 14/05/19. 3 July 2017.

12. Edge, J., et al., An alternative sensor-based method for glucose monitoring in children and young people with diabetes. Arch Dis Child, 2017. 102(6): p. 543–549.

13. Campbell, F.M., et al., Outcomes of using flash glucose monitoring technology by children and young people with type 1 diabetes in a single arm study. Pediatric Diabetes, 2018. 19(7): p. 1294–1301.

14. Landau, Z., et al., Use of flash glucose-sensing technology (FreeStyle Libre) in youth with type 1 diabetes: AWeSoMe study group real-life observational experience. Acta Diabetol, 2018. 55(12): p. 1303–1310.

15. Campbell, F. and J. Bolinder, FreeStyle Libre<sup>™</sup> Use for Self-Management of Diabetes in Teenagers and Young Adults. 2018. 67(Supplement 1): p. 158-LB.

16. McPhater, A., M. Gardiner, and G. Modgil, The impact of the Libre device for families and children with Type 1 diabetes. Diabetic Medicine, 2017. 34: p. 104–104.

17. Hellmund, R., R. Weitgasser, and D. Blissett, Cost calculation for a flash glucose monitoring system for UK adults with type 1 diabetes mellitus receiving intensive insulin treatment. Diabetes Res Clin Pract, 2018. 138: p. 193–200.

18. NHS Business Services Authority, June 2019 Drug Tariff. Available at: https://www.nhsbsa.nhs.uk/pharmacies-gp-practices-and-appliance-contractors/drug-tariff. Accessed 04/06/2019.

19. Leelarathna, L. and E.G. Wilmot, Flash forward: a review of flash glucose monitoring. Diabet Med, 2018. 35(4): p. 472–482.

20. RCPCH and H.P. Team., The case for investing in children and young people’s diabetes services. Available at: https://www.rcpch.ac.uk/resources/case-investing-children-young-peoples-diabetes-services. Accessed 14/05/19. 2019.

21. Hex, N., et al., Estimating the current and future costs of Type 1 and Type 2 diabetes in the UK, including direct health costs and indirect societal and productivity costs. Diabet Med, 2012. 29(7): p. 855–62.

22. Bristol North Somerset and South Gloucestershire Clinical Commissioning Group, Commissioning Policy Abbott FreeStyle Libre® Flash Glucose Monitoring System Criteria Based Access. Available at: https://bnssgccg-media.ams3.cdn.digitaloceanspaces.com/attachments/Freestyle_Libre_Policy_CBAv1920.1.00_.pdf. 2019.

23. Deidda, M., et al., A framework for conducting economic evaluations alongside natural experiments. Social science & medicine (1982), 2019. 220: p. 353–361.

24. Sutton, M., et al., Health Services and Delivery Research, in Economic analysis of service and delivery interventions in health care. 2018, NIHR Journals Library

25. Rovithis, D., Do health economic evaluations using observational data provide reliable assessment of treatment effects? Health economics review, 2013. 3(1): p. 21–21.

26. Rosenstock, I.M., V.J. Strecher, and M.H. Becker, Social learning theory and the Health Belief Model. Health Educ Q, 1988. 15(2): p. 175–83.

27. Murray, E., et al., Normalisation process theory: a framework for developing, evaluating and implementing complex interventions. BMC Med, 2010. 8: p. 63.

28. Braun, V. and V. Clarke, Using Thematic Anaysis in Psychology. QualResPsych, 2006. 3: p. 77–101.

29. NVivo qualitative data analysis software; Version 12 [program]. 2018.

30. NIHR INVOLVE, Guidance on co-producing a research project. Available at: https://www.invo.org.uk/wp-content/uploads/2019/04/Copro_Guidance_Feb19.pdf. Date accessed 16/07/2019. 2018.

